# Prevalence of Low Visual Acuity in children from public schools in Northeast of Brazil

**DOI:** 10.1101/2024.08.01.24311293

**Authors:** Lucas Neves de Oliveira, Matheus Gomes Reis Costa, Isadora Oliveira Santiago Pereira, Isabela Carolina Tokumoto, Joao Lucas de Magalhaes Leal Moreira, Matheus Carneiro Leal Freitas, Clarissa Silva Sampaio, Mateus Neves de Oliveira, Jose de Bessa Junior, Hermelino Lopes de Oliveira Neto

**Affiliations:** Division of Public Health, Department of Health, State University of Feira de Santana, Feira de Santana, Bahia, Brazil; Division of Public Health, Department of Health, Bahian School of Medicine and Public Health, Salvador, Bahia, Brazil; Division of Ophthalmology, Department of Health, State University of Feira de Santana, Feira de Santana, Bahia, Brazil

## Abstract

**OBJECTIVE:** To describe the prevalence of Low Visual Acuity (LVA) in public school students in Feira de Santana (FSA), Bahia (BA).

**METHODS:** This was an observational, cross-sectional, exploratory study. The sample consisted of schoolchildren from the 2nd to the 4th grade of five public schools in FSA/BA. Data collection was carried out in the schools themselves, with a sociodemographic and clinical questionnaire applied and Visual Acuity (VA) measured using the Snellen “E” optotype chart. LVA was defined as uncorrected VA < 20/25 in at least one eye.

**RESULTS:** The sample consisted of 358 children, with a median age of 9 [8-10] years, of which 189 (52.9%) were female. 248 (69.3%) individuals had never been to an ophthalmologist. LVA was found in 105 (29.3%) schoolchildren, and of these, 7.6% (8/105) current used glasses. Factors associated with LVA were female gender and white ethnicity. LVA was evidenced in 60 (31.7%) schoolchildren with excessive screen use and in 35 (25.5%) without excessive use (OR 1.35; 95% CI 0.83 - 2.19, p = 0.222), and excessive screen use was associated with visual signs/symptoms such as tearing and eye itching.

**CONCLUSION:** LVA was observed in approximately 30% of children in public schools in the interior of Bahia, and less than 10% of these current used glasses. Our study reinforces the importance of visual screening of schoolchildren through active search in our region and the creation of strategies to facilitate access to ophthalmological consultations and glasses.

## INTRODUCTION

Vision, among the five senses, is the most dominant and the primary means of integrating the individual with the external environment, with a large part of knowledge being acquired visually^1^. Visual problems impair learning and social interactions, compromising intellectual, academic, professional development, as well as communication and socialization skills^2^.

School-aged children are particularly affected by vision impairment. Initially, visual problems may not be perceived by the family, mainly due to the absence of signs or complaints. Over time, significant visual effort becomes evident in the teaching-learning process^3^. If persistent, these problems affect the child’s academic performance and socialization^4^.

According to the World Health Organization, there are approximately 1.4 million children with visual impairment worldwide, with 90% living in developing countries. Each year, 500,000 children become blind, and about 80% of childhood blindness causes are preventable or treatable^1^. It is estimated that the prevalence of childhood blindness in Brazil is 4/10,000 children^5^. Concerning reversible blindness, the leading cause of childhood blindness, uncorrected refractive errors are the primary causes of low vision in school-aged children^1^.

In Brazil, there is limited data on the prevalence of visual impairment in schoolchildren, and we are unaware of data in Bahia. Additionally, many studies are outdated, and most were conducted in the South and Southeast regions. A study conducted in Sorocaba, São Paulo, showed a prevalence of Low Visual Acuity (LVA) of 13.1% in public school children^6^. In Londrina, Paraná, the prevalence of LVA was demonstrated in 17.1% of public school students^7^. In Patos de Minas, Minas Gerais, the prevalence of visual impairment in schoolchildren was 20.9%^8^.

Early diagnosis of visual disorders has been suggested as a strategy to prevent future problems, including amblyopia, and alterations in neuropsychomotor and social development^9^. From a public health perspective, routine visual acuity assessment is essential for promoting eye health, contributing to reducing high levels of school dropout and poor academic performance ^6,10^.

Given the importance of early diagnosis, the scarcity of data in national literature, especially in the Northeast, and the absence of studies in Bahia, the aim of this study was to evaluate the prevalence of low visual acuity in public school children in Feira de Santana, Bahia.

## MATERIAL AND METHODS

### Study Design

This is an observational, cross-sectional, exploratory study. The sample consisted of elementary school students from the 2nd to the 4th grade, regularly enrolled in five municipal public schools located in Feira de Santana, Bahia. The schools are located in the city’s outskirts and were randomly selected by the Municipal Health Department. All students in the included grades were invited to participate. The evaluation period was from August 2022 to May 2023.

### Data Collection

Data were collected at the schools by medical students from the Visual Disorders Combat League (LCDV) of the State University of Feira de Santana (UEFS), adequately trained by ophthalmologists at an eye hospital. Initially, a sociodemographic and clinical questionnaire was administered to parents/guardians, including reports of recurring ophthalmic signs/symptoms and excessive screen use.

Visual Acuity (VA) was assessed using the Snellen “E” optotype chart, positioned 6 meters away and 1.5 meters high. The test was conducted without correction and, for children currently using glasses, it was repeated with optical correction ^7,11-13^.

### Definitions and Classifications

VA was classified as normal vision (VA ≥ 20/25 or 0.8), mild visual impairment (VA < 20/25 or 0.8 and ≥ 20/63 or 0.3), moderate visual impairment (VA < 20/63 or 0.3 and ≥ 20/160 or 0.125), and severe/profound visual impairment (VA < 20/160 or 0.125 and ≥ 20/1000 or 0.02)^13,14^. LVA was defined as uncorrected VA < 20/25 in at least one eye ^6-8,15^.

Excessive screen use was defined by a single question answered by parents. Screens included: cell phones, tablets, computers, and televisions.

### Statistical Analysis

Quantitative variables were described using measures of central tendency and dispersion. Qualitative variables were described in absolute values, percentages, and proportions. Categorical variables were compared using the chi-square test, and Odds Ratio was used as a measure of association for categorical variables. A p-value < 0.05 was considered statistically significant, and a 95% confidence interval was presented as a measure of precision. Data analysis was performed using IBM SPSS version 23.0, and graphs were created using GraphPad Prism version 10.2.2.

### Ethical Aspects

All children signed the Free and Informed Assent Term, and their guardians signed the Free and Informed Consent Term. All students identified with LVA were appropriately referred for a complete ophthalmologic consultation.

The activities developed are part of the project “A new look: a project to combat visual disorders in basic education,” approved by the Research Ethics Committee of the State University of Feira de Santana (UEFS), under CAAE: 56993722.5.0000.005. The project was executed by the Visual Disorders Combat League (LCDV) from UEFS in partnership with a specialized eye hospital located in Feira de Santana, Bahia.

## RESULTS

### Study Population

The sample consisted of 358 children, with 136 (38%) in the 2nd grade, 119 (33.2%) in the 3rd grade, and 103 (28.8%) in the 4th grade. The median age was 9 [8-10] years, and 189 (52.8%) were female. 311 (86.9%) self-identified as black/brown, and 241 (67.3%) had a family income ≤ 1 minimum wage. 248 (69.3%) students had never visited an ophthalmologist.

Thirty (8.4%) children reported previous or current use of glasses. Of these, 10 (33%) were current users, and the other 20 (67%) had stopped using them. The sample characteristics are detailed in Table 1.

**Table 1.** Baseline Characteristics of the Study Participants (N=358)

|  |  |
| --- | --- |
| Age (years) - median [interquartile range] | 9 [8-10] |
| Gender - n (%) |  |
| Male | 169 (47,2) |
| Female | 189 (52,8) |
| Grade - n (%) |  |
| 2nd | 136 (38) |
| 3rd | 119 (33,2) |
| 4th | 103 (28,8) |
| Race (self-reported) - n (%) |  |
| Black | 106 (29,6) |
| Brown | 205 (57,3) |
| White | 39 (10,9) |
| Yellow | 4 (1,1) |
| Indigenous | 4 (1,1) |
| Family income - n (%) |  |
| ≥ 3 minimum wage | 13 (3) |
| Between 1 and 3 minimum wage | 104 (29,1) |
| ≤ 1 minimum wage | 241 (67,3) |
| Residence - n (%) |  |
| Urban area | 331 (92,5) |
| Rural area | 27 (7,5) |
| Prematurity - n (%) |  |
| No | 323 (90,2) |
| 32 to 36 weeks | 30 (8,4) |
| 28 to 31 weeks | 4 (1,1) |
| ≤ 28 weeks | 1 (0,3) |
| Sistemic disease - n (%) |  |
| Total | 52 (14,5) |
| Respiratory diseases | 28 (7,8) |
| Rhinitis/sinusitis | 14 (3,9) |
| Asthma | 13 (3,6) |
| Previous consultation with an ophthalmologist - n (%) |  |
| No | 248 (69,3) |
| Yes | 110 (30,7) |
| Use of glasses - n (%) |  |
| Previous or current |  |
| No | 328 (91,6) |
| Yes | 30 (8,4) |
| Current |  |
| No | 348 (97,2) |
| Yes | 10 (2,8) |

### Visual Acuity

LVA (uncorrected visual acuity < 20/25 in at least one eye) was found in 105 (29.3%) students. Of these, 7.6% (8/105) were current glasses users, and after correcting Visual Acuity (VA) with glasses, 6 still had visual impairment in at least one eye. The distribution of VA is detailed in Table 2.

**Table 2.** Visual Acuity (VA) distribution (N=358)

| **Table 2. Visual Acuity (VA) distribution (N=358)** |  |  |
| --- | --- | --- |
| **VA Classification** | **Uncorrected VA n (%)** | **Current glasses users n (%)** |
| Normal vision ($\geq 20/25$) both eyes | 253 (70,7) | 2 (0,8) |
| Normal vision ($\geq 20/25$) one eye only | 29 (8,1) | 1 (3,4) |
| Mild visual impairment (< 20/25 e $\geq 20/63$) better eye | 56 (15,6) | 3 (5,3) |
| Moderate visual impairment (< 20/63 e $\geq 20/160$) better eye | 14 (3,9) | 2 (14,3) |
| Severe/profound visual impairment (< 20/160 e $\geq 20/1000$) better eye | 6 (1,7) | 2 (33,3) |
| Total | 358 (100) | 10 (2,8) |

In the univariate analysis, the variables associated to LVA were female gender (OR 2.12; 95% CI 1.32 - 3.41, p = 0.002) and white race (OR 2.57; 95% CI 1.31 - 5.05, p = 0.006). Age, family income, and prematurity were not associated (Table 3).

**Table 3.** Univariate analysis - variables associated to Low Visual Acuity (N = 358)

|  |  |  |  |
| --- | --- | --- | --- |
| **Variables** | **OR** | **CI 95%** | **P value** |
| Female | **2,12** | **1,32 - 3,41** | **0,002** |
| Age > 11 years | 2,47 | 0,85 - 7,24 | 0,098 |
| White race | **2,57** | **1,31 - 5,05** | **0,006** |
| Family income > 1 minimum wage | 1,25 | 0,77 - 2,02 | 0,362 |
| Prematurity | 1,12 | 0,53 - 2,37 | 0,774 |

### Vision and Excessive Screen Use

In the analysis of these aspects, our sample was reduced to 330 students due to incomplete data in this aspect for 28 subjects.

Excessive screen use was found in 189 (57.3%) children. LVA was observed in 60 (31.7%) students with excessive screen use and in 36 (25.5%) without excessive screen use (OR 1.35; 95% CI 0.83 - 2.19, p = 0.222).

The frequency of visual signs/symptoms in individuals with and without excessive screen use is shown in Graph 1. In the univariate analysis, there was an association between screen use and tearing (OR 1.62; 95% CI 1.02 - 2.56, p = 0.040), ocular pruritus (OR 1.73; 95% CI 1.11 - 2.71, p = 0.015), headache (OR 2.62; 95% CI 1.65 - 4.14, p < 0.001), photosensitivity (OR 2.92; 95% CI 1.83 - 4.66, p < 0.001), and blurred vision (OR 1.92; 95% CI 1.14 - 3.22, p = 0.014). We could not demonstrate an association with ocular hyperemia (OR 1.52; 95% CI 0.96 - 2.40, p = 0.076) and ocular pain (OR 1.37; 95% CI 0.85 - 2.22, p = 0.199), despite the higher prevalence in the group with excessive screen use.

**Graph 1:**
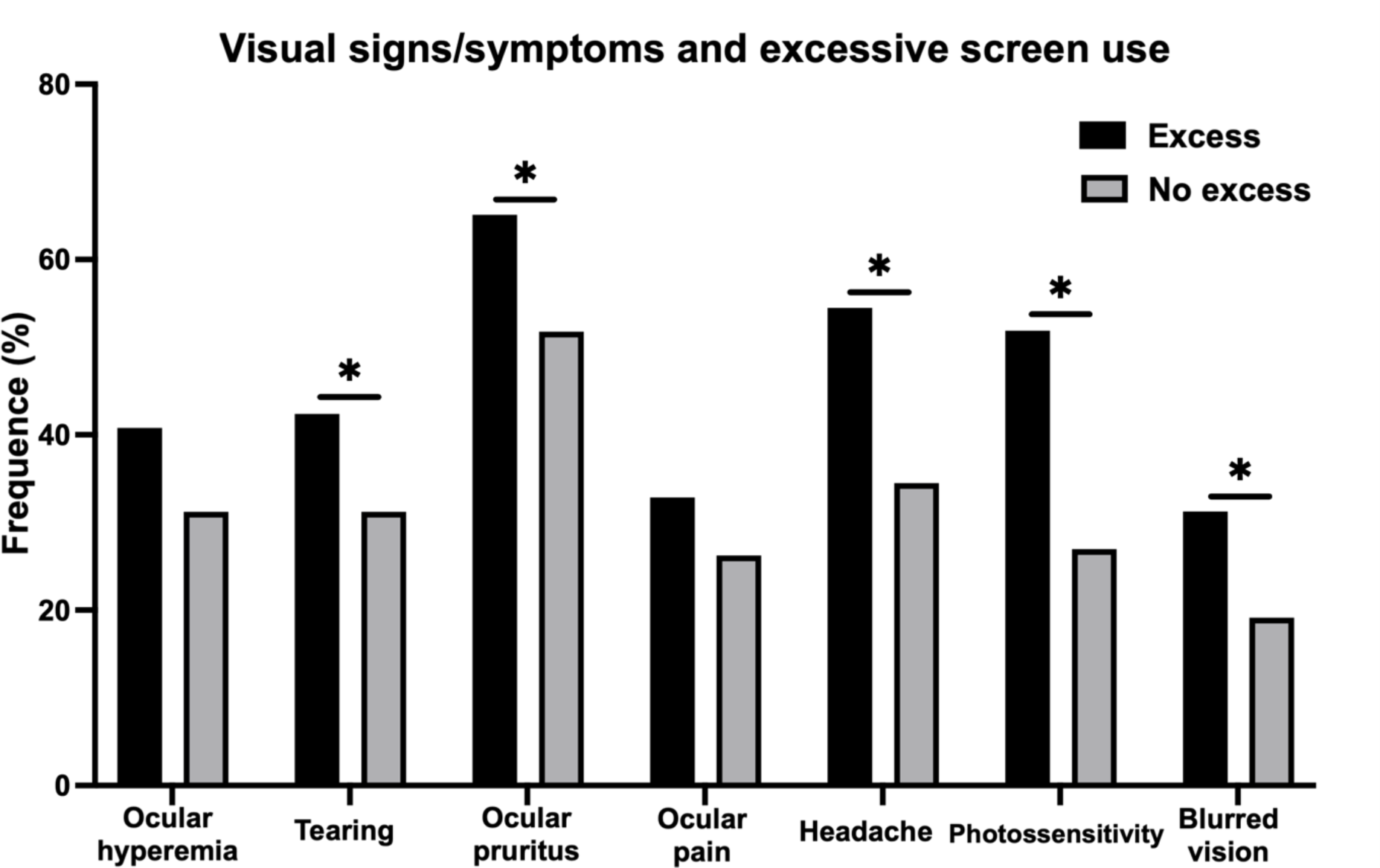
Visual sign/symptoms in schoolers with or without excessive screen use (N=330) *: p<0,05, X2.

## DISCUSSION

The prevalence of LVA in public school children in Feira de Santana, Bahia, was 29.3%. In national literature, the prevalence of LVA in schoolchildren ranged from 9% to 20.87%^6-8,11,13,15^. Our study found a higher prevalence of LVA than previous literature data. These differences may be explained by regional, methodological differences and sample characteristics.

The prevalence of current glasses use in this study was 2.8%. In previous studies in Brazil, the prevalence ranged from 2.4% to 4.5% ^6,7,11,13,15^. Regarding current glasses use in children with LVA, our study revealed a prevalence of 7.6%, while previous national studies showed a prevalence ranging from 10.52% to 40%^6,8,11,13^.

In addition to the higher prevalence of LVA in the studied population, a lower prevalence of current glasses use was demonstrated in children with low vision. It is noteworthy that approximately 70% of the students had never had an ophthalmological consultation. The Brazilian Society of Pediatric Ophthalmology recommends a complete routine ophthalmological consultation from six to twelve months of age and another consultation from three to five years of age, with the frequency of subsequent consultations being determined by the ophthalmologist, usually on an annual basis^16^. The findings reinforce the importance of conducting visual screening actions through active search in our region, as well as creating strategies to facilitate access to ophthalmological consultations and glasses.

The early detection and treatment of visual impairment in infants aim to ensure normal physical and cognitive development. Motor development and communication ability are impaired in infants with visual impairment because gestures and social behaviors are learned through visual feedback ^1,5^. There is an additional risk of developing amblyopia, characterized by low vision due to abnormal development of the visual cortex during childhood, which can affect one or both eyes^9,17^. Amblyopia should ideally be treated until ages 7-8. Some studies indicate benefit in treatment at older ages, but it is consensus that early correction provides the best prognosis^18^.

From a public health perspective, population investigation by ophthalmologists is unfeasible and costly, making routine visual screening by adequately trained non-medical personnel essential^5,19^.

A low-cost strategy capable of enabling large-scale visual screening is training teachers to apply the VA test using the Snellen chart. Other authors have recommended and validated this strategy^3,6,19,20^. Based on these premises, the Visual Disorders Combat League was created in 2021 by medical students from the State University of Feira de Santana. Grounded on the university triad and focusing on extension, one of league’s objectives is to promote eye health for children lacking ophthalmological care in Feira de Santana/Bahia and the surrounding region.

Factors associated with LVA in this study were female gender and white race. This association with female gender has been evidenced in other research^6,21^. No national studies associating ethnicity and LVA were found, but research in the United States showed that children who self-identified as black had worse VA^22,23^. Foreign literature also reports an association between family income and LVA^23,24^, a relationship not demonstrated in this study. Further national studies are needed to better elucidate these factors.

LVA was found more frequently in subjects with excessive screen use (31.7% vs 26.5%) compared to those without excessive use, although we did not find a statistically significant difference. However, we demonstrated an association between excessive screen use and some ophthalmological symptoms. There are still few studies on this topic in the literature, as the issue of screens is relatively recent, and the true impact on eye health is still unknown^25^.

A recent meta-analysis revealed that excessive smartphone use can increase the chance of ocular symptoms such as blurred vision, as well as myopia, asthenopia, and ocular surface diseases^26^. Besides the neuropsychomotor and social benefit, restricting prolonged screen use seems to positively impact eye health, making parental involvement indispensable in monitoring and regulating excess^26-28^.

This study has some limitations. In 2021, it was estimated that Feira de Santana had 16,364 children enrolled in the 2nd to 4th grade in municipal schools^29^. Due to logistical difficulties in screening and team size limitations, we had a relatively small (N=358) and non-probabilistic sample size. Additionally, the study’s unicentric nature and the subjective method of defining screen time limit our external validity. Despite these limitations, this study is pioneering in our region and presents relevant data for public health.

## CONCLUSION

Low visual acuity was observed in approximately 30% of public school children in the interior of Bahia, and less than 10% of these were current glasses users. About 70% of the children had never seen an ophthalmologist. Excessive screen use seems to be a significant issue and may harm eye health. Our study reinforces the importance of active visual screening of schoolchildren in our region, as well as creating strategies to facilitate access to ophthalmological consultations and glasses.

## Data Availability

All data produced in the present work are contained in the manuscript

